# Genetic variation in activating clopidogrel: longer-term outcomes in a large community cohort

**DOI:** 10.1101/2021.04.19.21255559

**Authors:** Luke C. Pilling, Deniz Türkmen, Hannah Fullalove, Janice L. Atkins, João Delgado, Chia-Ling Kuo, George A. Kuchel, Luigi Ferrucci, Jack Bowden, Jane A.H. Masoli, David Melzer

**Affiliations:** Epidemiology and Public Health Group, College of Medicine and Health, University of Exeter, Exeter, UK; UConn Center on Aging, University of Connecticut, Farmington, CT, USA; Connecticut Convergence Institute for Translation in Regenerative Engineering, University of Connecticut, CT, USA; National Institute on Aging, Baltimore, MD, USA; Exeter Diabetes Group (ExCEED), College of Medicine and Health, University of Exeter, Exeter, UK; Department of Healthcare for Older People, Royal Devon and Exeter Hospital, Barrack Road, Exeter, UK

## Abstract

**Background:** The antiplatelet drug clopidogrel is commonly prescribed for stroke and myocardial infarction (MI) prevention. Clopidogrel prodrug is predominantly activated by liver enzyme CYP2C19. *CYP2C19* Loss-of-function (LoF) genetic variants have been linked to excess morbidity mainly in patients hospitalized for acute ischemic events and related interventions. Little is known about the magnitude of impact of LoF variants in family practice, especially over long periods of exposure. We aimed to determine whether *CYP2C19* LoF alleles increase risk of ischemic stroke and MI in primary care patients prescribed clopidogrel for up to 18 years.

**Methods:** Retrospective cohort analysis of 7,483 European-ancestry adults from the UK Biobank study with genetic and linked primary care data, aged 36 to 79 years at first clopidogrel prescription. We examined *CYP2C19* LoF variant (*2-*8) associations with incident hospital-diagnosed ischemic stroke and MI in patients prescribed clopidogrel for at least 2 months using time-to-event models, with secondary analysis of the *17 gain of function variant.

**Results:** 28.7% (n=2,144/7,483) of included subjects (mean age 63 years at first clopidogrel prescription) carried at least one C*YP2C19* intermediate or low metabolizer LoF variant. 1.9% of LoF variant carriers had an incident ischemic stroke whilst prescribed clopidogrel (mean 2.6 years, range 2 months to 18 years), versus 1.3% without the variants (0.6% absolute excess: Hazard Ratio 1.53: 95% CI 1.04 to 2.26, p=0.031). Additionally, 26.4% of C*YP2C19* LoF variant carriers had an incident MI versus 24.1% (HR 1.14: 1.04 to 1.26, p=.008). Adjustment for aspirin co-prescription produced similar estimates. In lifetables using observed incidence rates, 22.5% (95% CI 14.4% to 34.0%) of *CYP2C19* LoF carriers on clopidogrel were projected to develop an ischemic stroke by age 79 (the oldest age in the study), compared with 15.4% (95% CI 11.4% to 20.5%) in non-carriers: the absolute excess stroke incidence with LoF variants was 7.1% by age 79.

**Conclusion:** In family practice patients on clopidogrel, *CYP2C19* LoF variants are associated with substantially higher incidence of ischemic events. Genotype-guided (or routine) prescription of antiplatelet medications unaffected by *CYP2C19* variants may improve outcomes in patients for whom clopidogrel is currently indicated.

## Introduction

Platelet aggregation, the process by which platelets adhere to each other, has long been recognized as critical for hemostatic plug formation and thrombosis (1). Antiplatelet therapy that reduces platelet aggregation has become a central part of treatment to reduce cardiovascular and cerebrovascular disease incidence in at-risk patient groups (2). Clopidogrel is an irreversible antagonist for the platelet P2Y_12_ adenosine diphosphate receptor, thereby inhibiting platelet function and reducing likelihood of thrombosis (3). In 2018 it was the 39^th^ most commonly prescribed drug in the USA (20 million prescriptions) (4). Clopidogrel is a prodrug that requires transformation to its bioactive form by cytochrome P-450 (CYP) enzymes, primarily CYP2C19 (3). Loss-of-function (LoF) genetic polymorphisms in *CYP2C19* impair clopidogrel metabolism, with carriers of any LoF variant reported to have a 32.4% reduction in plasma active metabolite levels compared to non-carriers (3). Clopidogrel resistance (high on-treatment platelet reactivity) is predominantly caused by *CYP2C19* LoF variants, with a recent study of clopidogrel on-treatment reactivity reporting that 71.7% of LoF carriers show clopidogrel resistance, compared to 32.1% of non-carriers (5). *CYP2C19* LoF carriers therefore have decreased platelet inhibition with consequently increased risk of thrombosis (6).

LoF variants in the *CYP2C19* gene are common in many populations, with the *2 allele ranging from 15% frequency in Europeans (with ∼27% of the population carrying at least one copy) to >30% in Asian populations (7). A recent 2017 meta-analysis of 15 studies (12 East Asian, 3 European ancestry predominantly from the USA) included 4,762 stroke or transient ischemic attack (TIA) patients treated with clopidogrel; patients carrying any *CYP2C19* LoF alleles had nearly double the rates of incident strokes (Risk Ratio [RR] 1.92: 95% Confidence Intervals 1.10 to 2.06, p=0.01) compared to non-carriers (8). However, most studies thus far focused on specific hospitalized patient groups with short (up to 1 year) follow-ups. A 2011 systematic review concluded that a significant association with stent thrombosis was due to small study effect bias and replication diversity (9). Although the US Food and Drug Administration added a Boxed Warning to the clopidogrel label regarding poor metabolizers due to *CYP2C19* variants (10) in 2010, there is continued debate about whether the magnitude of effect of these variants on clinical outcomes justifies *CYP2C19* genotype (6) guided prescription or use of alternative medications unaffected by these variants.

Given the widespread continuing use of clopidogrel, and the current scarcity of evidence on clinical outcomes due to the LoF variants in the primary care setting, we aimed to estimate the association between *CYP2C19* LoF alleles and incident hospital-diagnosed ischemic stroke and MI in UK Biobank participants who were prescribed clopidogrel. We also aimed to estimate longer-term (>1 year) outcomes, as these have previously been understudied, and to estimate outcomes of non-*CYP2C19* variants reported to have similar effects (11).

## Methods

### UK Biobank cohort

The UK Biobank recruited 503,325 community-based volunteers aged 40-70 years who visited one of 22 assessment centers in England, Scotland or Wales between 2006 and 2010 (12). Data collected at the baseline assessment included extensive questionnaires on demographic, health, and lifestyle information. Anthropometric measures were also taken, in addition to blood samples for future biochemical and genetic analysis. Ethical approval for the UK Biobank study was obtained from the North West Multi-Centre Research Ethics Committee. This research was conducted under UK Biobank application 14631 (PI: DM). UK Biobank volunteers tended to be healthier than the general population at baseline assessment (13), however this largely prospective analysis of linked primary care data is less affected by sample response patterns than baseline parameters.

### Prescription data

General practice prescriptions data is currently available for 230,096 (45.7%) participants, with >57 million prescribing events recorded. UK Biobank obtained data from the three main primary care electronic clinical record data providers (England: TPP/Vision; Scotland and Wales: EMIS/Vision). The extract date differs between the supplier’s computer systems, and we used the following censoring dates: June 2016 (England), April 2017 (Scotland), and September 2017 (Wales). Detailed description of the data extracted and limitations are available from UK Biobank (14). Data includes prescription dates, drug code (in clinical Read v2, British National Formulary (BNF), or Dictionary of Medicines and Devices format (depending on supplier), drug name, and quantity, where available. We identified all prescribing events for clopidogrel and aspirin. Details for drug names and clinical codes used are available in the Supplementary Information and Supplementary Table 1.

In the prescribing data we identified all records matching a clopidogrel code and for each patient determined the date first prescribed, date last prescribed, number of total prescriptions, and average number of prescriptions over the time period. We analyzed data for patients who received two or more prescriptions, with at least one clopidogrel prescription for each 2-month period between the dates first/last prescribed. This last criterion excluded participants with longer gaps between prescriptions, who may not have been taking clopidogrel for the entire study period, thus minimizing immortal time bias (15).

Participants never prescribed clopidogrel were used for sensitivity analysis of *CYP2C19* LoF variants and relevant vascular outcomes.

### *CYP2C19* genetic variants

Directly genotyped genetic variants (n=805,426) were available for 488,377 UK Biobank participants, based on two almost identical genotyping platforms sharing >95% of variants: the Affymetrix Axiom UKB array (in 438,427 participants) and the Affymetrix UKBiLEVE array (in 49,950 participants). After extensive quality control by the central UK Biobank team (16), genotype imputation was successful in 487,442 participants and increased the number of genetic variants to ∼96 million (16). Our primary analysis included 451,367 participants (93%) identified as genetically European (identified by genetic clustering, as described previously (17)), to reduce bias in genetic studies from population stratification, which arises when different ancestral groups are analyzed together in genetic analyses (18). Secondary analysis in other ancestral groups was not possible due to low numbers in the clopidogrel patients (n=492 non-Europeans with sufficient clopidogrel data; this group included all other ancestries such as South or East Asian, African, and mixed).

We analyzed the *CYP2C19* genetic variants (*2-*8 and *17) with well-documented effects on clopidogrel metabolism in the literature (9) and in the PharmGKB database (11): three loss-of-function alleles (*2, *3 and *8; intermediate or poor metabolizers) and one gain-of-function allele (*17; rapid metabolizer) were either directly genotyped (*3 and *8) or imputed with high confidence (*2 and *17: >99.9% imputation confidence). *CYP2C19*\*4 was imputed with <80% imputation confidence (74.4%) so was excluded from analysis (*4 is very rare so this has minimal impact on the analysis: with 0.25% allele frequency reported in the GnomAD (7) database https://gnomad.broadinstitute.org/). *CYP2C19* *5, *6 and *7 were not available in the genotyping data, as they are extremely rare, especially in European populations (<0.1% allele frequency in GnomAD (7)). See Supplementary Table 2 for details on individual variants used.

### Non-*CYP2C19* genetic variants

We also investigated non-*CYP2C19* genetic variants that affect clopidogrel from two sources: we searched the PharmGKB database (11) for variants classified as being supported by moderate or high clinical annotation levels of evidence, i.e. associations that replicated in subsequent studies, even if the original study was small. In addition to variants in *CYP2C19* (level 1 “high” levels of evidence) one variant in *CES1* has level 2B “moderate” evidence: rs71647871 was directly genotyped on the microarray but is rare, with only 249 heterozygotes in the clopidogrel analysis group (no homozygotes).

Lastly, we investigated variants identified in a genome-wide association study of clopidogrel active metabolite levels (rs187941554 and rs80343429) (19). Both variants are available in the UK Biobank imputed genotype data: rs187941554 was imputed with high confidence (99.9%), however the overall rs80343429 imputation accuracy was 81.7% (n=3 individual participants with low-confidence genotype calls - imputed genotype dose >0.25 and <0.75 - were excluded from analysis).

### Disease ascertainment

Incident ischemic stroke and MI ascertainment from hospital admissions records (Hospital Episode Statistics, HES) were available up to 14 years follow-up after baseline assessment, covering the entire period up to the date of censoring of primary care prescribing data (HES in England up to 30 September 2020: data from Scotland and Wales censored to 31 August 2020 and 28 Feb 2018, respectively). Diagnosis of ischemic stroke was ascertained using ICD-10 code I63*. Diagnosis of MI was ascertained using ICD-10 codes I21*; I22*; I23*; I24*; I25*.

For prevalent ascertainment of diagnoses at baseline of transient ischemic attack (TIA), MI/angina, atrial fibrillation, hypertension or peripheral vascular disease we used primary care records. Diagnosis codes (read v2) were downloaded from the CALIBER phenotype library (https://portal.caliberresearch.org/disease-or-syndrome) (20). Corresponding CTV3 codes were identified using the UK Biobank “Clinical coding classification systems and maps” resource (https://biobank.ctsu.ox.ac.uk/crystal/refer.cgi?id=592).

### Statistical analysis

Data are from multiple sources (demographic and genetic data from UK Biobank assessment, clopidogrel prescribing data from linked electronic primary care records, and stroke and MI outcomes from linked electronic hospital inpatient records). Patients with any missing data needed for analysis were excluded, with details shown in the results.

Estimates of associations between genotype and incident disease were obtained from Cox’s proportions hazards regression models, with adjustment for age at first prescription, sex, and the first ten principal components of genetic ancestry (to control for population substructure). Patients entered the model 7 days after the date of first prescription of clopidogrel; this was to exclude the small number of events that occurred immediately after being prescribed clopidogrel, before it could reasonably have had a preventative effect. Seven days was chosen because the effect of clopidogrel on platelet aggregation was significantly different between *CYP2C19* genotype groups after 7-10 days in a study of 375 patients (21). Patients exited the model on the last known date of clopidogrel prescription. STATA (v15.1) software was used for analysis. We visually inspected Kaplan-Meier plots and used STATA function ‘estat phtest, detail’ to generate Schoenfeld residuals to test for violations of the proportional-hazards assumption.

Population attributable fraction in this analysis is the estimated proportion of all cases that would not have occurred if no participants carried the clopidogrel modifying genetic variants. It was calculated by multiplying the proportion of incident ischemic strokes in the genotype carriers by (1 – 1 / Hazard Ratio for ischemic stroke) and similarly for MI (22). This method is preferred to simple observed vs. expected cases as it uses the estimate from the time-to-event model, which has been adjusted for covariates (22). Lifetable probabilities of incident outcomes were estimated from age 37 to 79 (the minimum and maximum ages the patients were first prescribed clopidogrel, respectively) by *CYP2C19* genotype, applying observed incidence rates in each age group to a notional cohort (where all participants in the analysis had reached the maximum age group), estimating cumulative incident case numbers, using STATA command ‘sts list’.

### Patient and public involvement

Patients and participants were and are extensively involved in the UK Biobank study itself. No patients were involved in developing the research question or the outcomes tested. A lay abstract was reviewed by members of the Peninsula Public Engagement Group (“PenPEG”) in Exeter, who provided positive feedback on the importance of the study from a patient perspective.

## Results

7,483 UK Biobank participants of European ancestry met inclusion criteria, with at least two prescriptions of clopidogrel recorded in primary care data (see Figure 1 for cohort flowchart with detailed inclusion/exclusion criteria). The mean age at first prescription was 64.3 years (SD 7.2), and 2,563 were female (34.2%). The clopidogrel exposure period ranged from 2 months to 18 years (mean 2.59 years, SD 2.98). Cohort selection is described in Table 1. Patients carrying at least one *CYP2C19* LoF variant (*2-*8, intermediate/poor clopidogrel metabolizers) did not have significantly different likelihood of receiving clopidogrel compared to non-carriers (Odds Ratio 1.01: 95% Confidence Intervals 0.96 to 1.05, p=0.75) and had similar prevalence of cardiovascular co-morbidities prior to diagnosis (Table 1).

**Figure 1:**
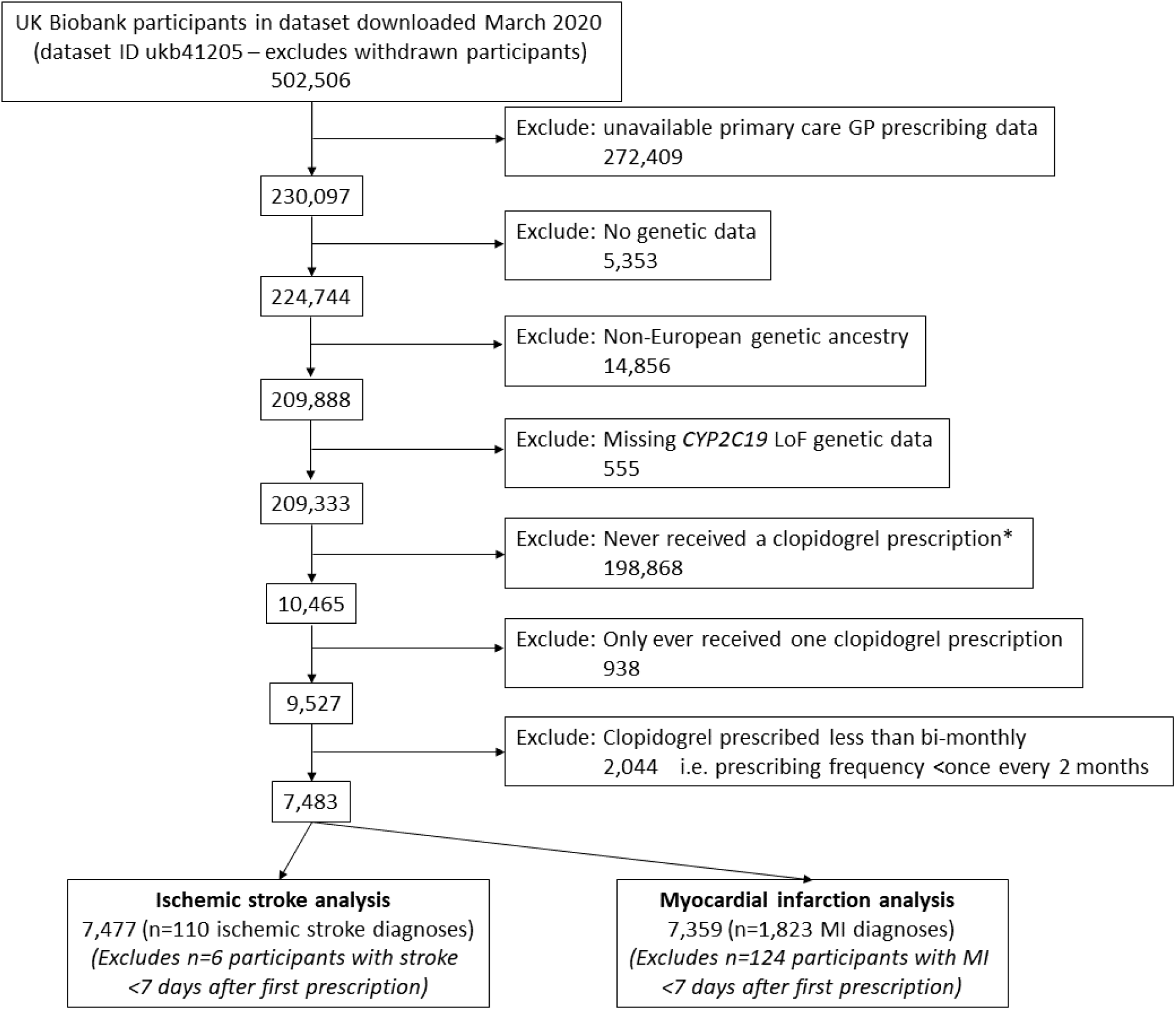
UK Biobank participants eligible for analysis: cohort flowchart. Cohort flowchart shows how the eligible participants to study were identified. Summary statistics for the 7,483 participants are available in Table 1. *Participants never prescribed clopidogrel were used for sensitivity analysis of CYP2C19 LoF variants and vascular outcomes.

**Table 1:** Summary of UK Biobank participants with GP-prescribed clopidogrel.

|  | **CYP2C19 genotype** |  |
| --- | --- | --- |
|  | **Normal metabolizer**<br>**(\*1/\*1)** | **Intermediate/poor**<br>**(any \*2-\*8)** |
| Number of participants (% of total n=7,483) | 5,338 (71.3) | 2,145 (28.7) |
| Females, n (% of genotype group) | 1,847 (34.6) | 712 (33.2) |
| Age at first clopidogrel prescription |  |  |
| Minimum : maximum | 36.5 to 78.8 | 37.6 to 78.1 |
| Mean (SD) | 64.1 (7.3) | 64.3 (7.2) |
| Diagnoses* prior to clopidogrel prescription, n (%) |  |  |
| Ischemic stroke | 509 (9.5) | 240 (11.2) |
| Transient ischemic attack | 1,142 (21.4) | 422 (19.7) |
| Myocardial infarction / angina | 2,980 (55.8) | 1,267 (59.1) |
| Atrial fibrillation | 408 (7.6) | 180 (8.4) |
| Hypertension | 3,072 (57.6) | 1,277 (59.5) |
| Peripheral vascular disease | 442 (8.3) | 177 (8.3) |
| Any of the above six diagnoses | 4,863 (91.1) | 1,982 (92.4) |
| Number of clopidogrel prescriptions |  |  |
| Minimum : maximum | 2 to 467.0 | 2 to 494.0 |
| Mean (SD) | 28.3 (36.7) | 27.9 (36.9) |
| Years between first and last clopidogrel prescription |  |  |
| Minimum : maximum | 0.2 to 17.9 | 0.2 to 17.3 |
| Mean (SD) | 2.6 (3.0) | 2.5 (2.9) |
| Also prescribed aspirin during clopidogrel exposure period, n (%) | 2,632 (49.3) | 1,119 (52.2) |
| Ischemic stroke during clopidogrel prescribing period, n (%) | 69 (1.3) | 41 (1.9) |
| Myocardial infarction during clopidogrel prescribing period, n (%) | 1,268 (24.1) | 554 (26.4) |

*Analysis of European-ancestry participants with >1 clopidogrel prescription in the available GP prescribing data. Participants excluded if clopidogrel prescribing frequency was less than once every 2 months. Events occurring after the last known date of clopidogrel prescription are also excluded. \* Ischemic stroke diagnoses are from hospital in-patient records only; all other disease also include diagnoses recorded in primary care.*

### *CYP2C19* LoF associations with incident ischemic stroke

Incident hospital-diagnosed ischemic stroke (cerebral infarction) was identified in 110 of 7,477 participants (1.47%) eligible for analysis (after excluding n=6 who had stroke within 7 days of first prescription). *CYP2C19* LoF (*2-*8, intermediate/poor metabolizers) carriers (n=2,144, 28.7%) were more likely to have an incident ischemic stroke whilst being prescribed clopidogrel than patients without *CYP2C19* LoF genetic variants (Hazard Ratio (HR) 1.53: 95% CI 1.04 to 2.26, p=0.031). See Table 2 for details and Figure 2A for Kaplan-Meier plot. 1.29% of the patients without any *CYP2C19* LoF genetic variants had an incident stroke during the prescribing period (mean 2.59 years between first and last clopidogrel prescription, SD 2.98) compared to 1.91% of the *CYP2C19* LoF carriers: this absolute 0.64% excess is statistically significant (95% CI 0.021% to 1.35%, p=0.027).

**Table 2:**
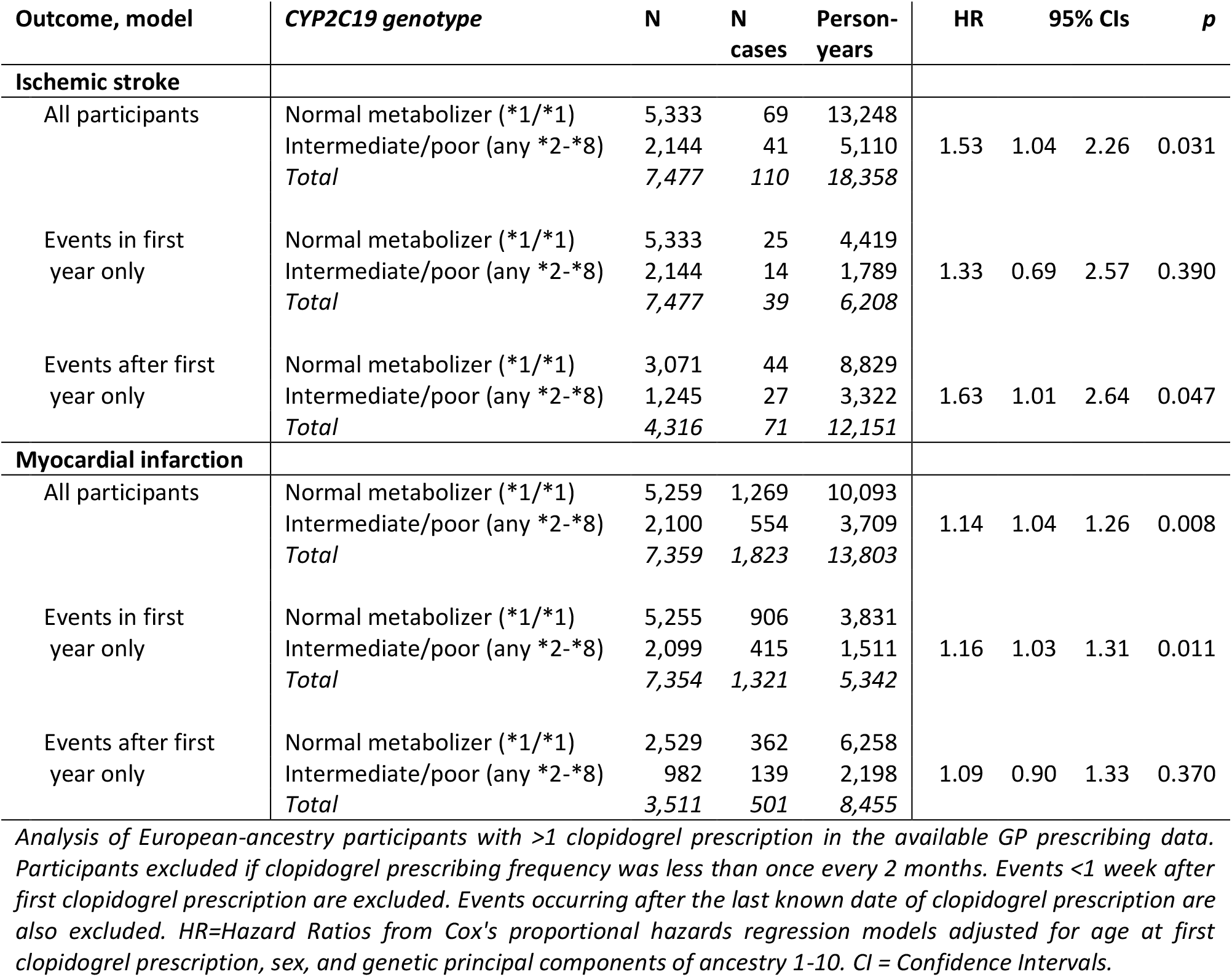
*CYP2C19* genotype associations with incident ischemic stroke and MI events in patients prescribed clopidogrel.

**Figure 2:**
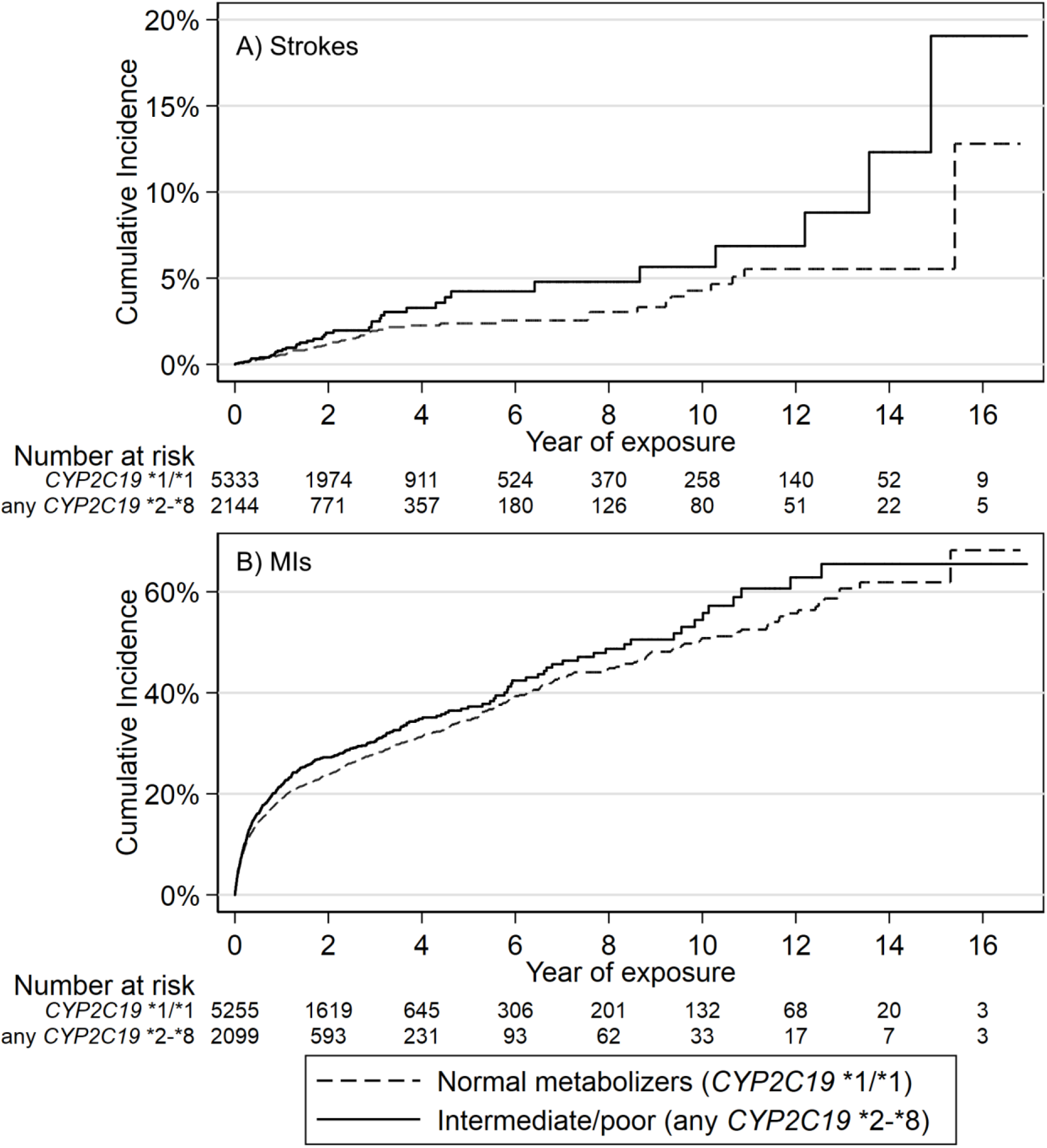
Kaplan-Meier plots of strokes and MIs in patients prescribed clopidogrel, stratified by *CYP2C19* Loss of Function genotypes. Kaplan-Meier failure plots for A) incident ischemic stroke and B) incident MI showing cumulative hazard (%) with increasing time taking clopidogrel in patients prescribed clopidogrel for at least 2 months, stratified by CYP2C19 genotype (carriers of any *2-*8 Loss of Function variant, “intermediate/poor metabolizers of clopidogrel,” vs. *1/*1 normal metabolizers).

To test whether the effect of *CYP2C19* LoF genotypes on ischemic stroke risk was specific to clopidogrel, and not via a separate but unknown biological mechanism, we analyzed the 198,868 UK Biobank participants with GP prescribing data in whom clopidogrel was never prescribed (see Figure 1). There were 512 incident ischemic stroke events in this group after baseline assessment and before the end of Feb 2016 (earliest censoring date for GP prescribing data). *CYP2C19* LoF carriers were not at significantly increased risk of ischemic strokes compared to non-carriers (HR 1.12: 95% CI 0.93 to 1.35, p=0.24).

To calculate the population attributable fraction (PAF) we multiplied the proportion of incident ischemic strokes that occurred in that patients who had intermediate or low metabolizer variants (41/110) by (1-1/HR for ischemic stroke = 1-1/1.53 = 0.346) giving 0.129 (see Methods). We therefore estimate the PAF for ischemic stroke - the proportion of ischemic strokes in this population that would not have occurred if clopidogrel efficacy was not affected by *CYP2C19* LoF variants - to be 12.9% (95% CI 1.4% to 20.8%). Kaplan-Meier curves for diagnosis of incident ischemic strokes in participants prescribed clopidogrel are shown in Figure 2. In lifetable estimates based on observed incidence rates from 36 to 79 (the minimum and maximum ages at which clopidogrel was first prescribed) the risk of ischemic stroke in CYP2C19 LoF carriers was 22.5% (95% CI 14.4% to 34.0%), compared with 15.4% (95% CI 11.4% to 20.5%) in non-carriers.

We performed secondary analyses estimating outcomes for participants prescribed clopidogrel for >1 year (n=4,316: 57.8%). *CYP2C19* LoF carriers had significantly increased risk of incident strokes (HR 1.63: 95% CI 1.01 to 2.64, p=0.047) in this period (Table 2). However the association with only those events within the first year of prescription was not statistically significant (HR 1.33: 95% CI 0.69 to 2.57, p=0.390). In tests of the proportional-hazards assumption (see Methods) in the model using all available events there was no evidence that the hazard ratio changed over time (p=0.42). The analysis using all participants suggests LoF carriers are at increased risk for the duration of the clopidogrel prescribing (HR 1.53: 95% CI 1.04 to 2.26, p=0.031).

We investigated hemorrhagic strokes (ICD-10 codes I61* and I62*), however there were too few diagnoses made (n=9 in the 7,483 participants included in the study) during the clopidogrel prescribing period, so no analysis was performed.

We also analyzed outcomes for the *17 (gain of function) genotype carriers (rapid metabolizers). *17 carriers were not at altered risk of incident stroke during the prescribing period (HR 0.98: 95% CI 0.61 to 1.57, p=0.930, Supplementary Table 3) compared to *1/*1 patients.

### *CYP2C19* LoF associations with incident myocardial infarction

Incident hospital-diagnosed myocardial infarctions (MI) occurred in 1,823 of 7,359 participants (24.8%), after excluding 124 subjects who had MIs within 7 days of first clopidogrel prescription. *CYP2C19* LoF (*2-*8, intermediate/poor metabolizers) carriers (n=2,100, 28.5%) were more likely to have an incident MI whilst being prescribed clopidogrel than patients without *CYP2C19* LoF genetic variants (HR 1.14: 95% CI 1.04 to 1.26, p=0.008, see Table 2 for details and Figure 2B for Kaplan-Meier plot). The PAF for MI was 3.73% (95% CI 1.2% to 6.3%).

We performed secondary analysis investigating the participants prescribed clopidogrel for >1 year (n=3,511: 47.7%). *CYP2C19* LoF carriers were not at significantly increased risk of incident MIs (HR 1.09: 95% CI 0.90 to 1.33, p=0.37) in this period (Table 2). However, LoF carriers did have significantly increased risk of MI within the first year of clopidogrel prescription (HR 1.16: 95% CI 1.03 to 1.31, p=0.011). In tests of the proportional-hazards assumption (see Methods) in the model using all available events there was no evidence that the hazard ratio changed over time (p=0.70). The analysis using all participants suggests LoF carriers are at increased risk for the duration of the clopidogrel prescribing (HR 1.14: 95% CI 1.04 to 1.26, p=0.008).

We also analyzed the *17 (gain of function) genotype carriers (rapid metabolizers). *17 carriers did not have significantly different risk of incident MI during the prescribing period (HR 0.95: 95% CI 0.85 to 1.06, p=0.33, Supplementary Table 3) compared to *1/*1 patients.

The association between *CYP2C19* LoF carriers and MI in participants never prescribed clopidogrel (n=4,390 MIs) was not significant (HR 1.03: 95% CI 0.96 to 1.10, p=0.41).

In secondary analysis of any ischemic event (stroke or MI, *n*=1,886 events in 7,354 patients included in analysis) *CYP2C19* LoF (*2-*8, intermediate/poor metabolizers) carriers were more likely to have an event whilst being prescribed clopidogrel than patients without *CYP2C19* LoF genetic variants (HR 1.17: 95% Cis 1.06 to 1.29, p=0.002).

### Dual antiplatelet therapy: clopidogrel co-prescribing with aspirin

Of 7,483 patients prescribed clopidogrel 3,730 (49.9%) were also prescribed aspirin at least once during the analysis period. *CYP2C19* LoF carriers had slightly higher likelihood of ever receiving a prescription of aspirin compared to non-carriers during their clopidogrel exposure period (n=1,118 (52.2%) vs. n=2,629 (49.3%), odds ratio (OR) from logistic regression 1.14: 95% CI 1.02 to 1.26, p=0.017). Ever being prescribed aspirin did not significantly interact with *CYP2C19* LoF carrier status in the model of incident stroke or MI (p>0.05). The association between *CYP2C19* LoF carrier status and incident stroke or MI was consistent (albeit attenuated due reduced sample sizes in subgroups) in participants who received an aspirin prescription (n=3,730. Stroke HR 1.61: 95% CI 0.87 to 2.98, p=0.13; MI HR 1.10; 95% CI 0.98 to 1.24, p=0.10) and those who did not (n=3,753. Stroke HR 1.48: 95% CI 0.89 to 2.46, p=0.13; MI HR 1.19: 95% CI 0.98 to 1.45, p=0.087).

### Analysis of non-*CYP2C19* genetic variation associated with clopidogrel active metabolite levels

A rare variant in *CES1* (rs71647871, n=249 CT heterozygotes) with moderate clinical annotation evidence for clopidogrel on the PharmGKB database (11) was not significantly associated with strokes or MIs (stroke HR 0.86: 95% CI 0.27 to 2.70, p=0.79; MI HR 1.26: 95% CI 0.99 to 1.60, p=0.057), although the MI association was borderline.

Two genetic variants identified in a GWAS of clopidogrel active metabolite levels (19) were also analyzed. Despite its low frequency in the population, rs187941554 (n=32 GA heterozygotes, 0.4% of 7,468) was associated with substantially raised risk of incident stroke (n strokes=2; HR 5.44: 95% CI 1.33 to 22.30, p=0.019) but not MIs (HR 1.20: 95% CI 0.62 to 2.32, p=0.58). The rs80343429 A allele was not associated with increased risk of stroke or MI (stroke HR 1.19: 95% CI 0.68 to 2.09, p=0.53; MI HR 1.09: 95% CI 0.94 to 1.26, p=0.25).

## Discussion

Clopidogrel requires metabolic activation to be effective, mainly by the CYP2C19 liver enzyme. *CYP2C19* LoF genetic variants (implying intermediate or low metabolizer status) have been shown to increase stroke and MI rates mainly in hospitalized patients with acute ischemic conditions (or undergoing related interventions such as cardiac stents (23)), with little data on outcomes in primary care or over longer follow-up periods. In this study of 7,483 UK Biobank primary care patients prescribed clopidogrel, carriers of *CYP2C19* LoF alleles had substantially increased risks of incident ischemic strokes and myocardial infarction. We also showed that carriers of LoF alleles are at increased risk of stroke even during longer-term (>1 year) therapy; the effect is not limited to acute stroke patients.

Our results are consistent with previous studies of clopidogrel efficacy in *CYP2C19* LoF genotype carriers, although the previous evidence was predominantly from high-risk acutely hospitalized patient groups (24–27), with limited follow-up periods (rarely over 12 months). Some studies reported that the effect of *CYP2C19* genotype on reducing clopidogrel efficacy was limited to the short-term (1-6 months) only (28,29), but in our community based longer term follow-up we found no evidence for deviation from the proportional hazards assumptions, with survival analysis results suggesting sustained effects over the follow-up period. We had limited sample size and therefore limited statistical power to estimate effects for specific periods, but the intermediate or low metabolizer variant association with excess incident strokes was significant in the subgroup prescribed clopidogrel for more than one year.

A recent 2019 meta-analysis of 16 randomized clinical trials (30) found that dual antiplatelet therapy with aspirin was associated with significantly lower stroke rates (RR 0.80: 0.72 to 0.89), however some evidence of increased major bleeding was also observed (RR 1.90: 1.33 to 2.72). The authors noted that common *CYP2C19* LoF variants may account for the effectiveness of dual therapy over clopidogrel monotherapy (30). In our study we found LoF carriers still had increased stroke or MI risk, even if co-prescribed aspirin.

Clopidogrel is a recommended antiplatelet medication for the prevention of stroke in the USA (2) and is the currently recommended antiplatelet medication for prevention of stroke or TIA in the UK (31). However, antiplatelet medications such as ticagrelor and prasugrel are not dependent on CYP2C19 metabolism and reportedly have better clinical outcomes in LoF carriers compared to clopidogrel (32). A recent meta-analysis of 16 studies (8 RCTs and 8 cohorts) concluded that genotype-guided antiplatelet treatment (primarily based on *CYP2C19* LoF alleles) improved patient outcomes for major cardiovascular events (e.g. Relative Risk for MI 0.45: 95% CI 0.35 to 0.58, p<0.00001), with decreased risk of major bleeding (27). Treatment strategy in the different metabolizer groups varied between individual studies, but included use of higher doses of clopidogrel (150 mg/d instead of the usual 75 mg/d), or prescribing of prasugrel or ticagrelor, or a combination. Other recent studies in different populations, especially in patients with acute coronary syndrome, also support this conclusion (24–26). Therefore, further work is needed in primary care populations to inform optimal antiplatelet care, given our estimate that over 12% of incident strokes in the UK study population on clopidogrel could be avoided if low and intermediate metabolizers could achieve the same clinical outcomes as are being achieved in the normal metabolizer group. Economic analysis in the UK comparing clopidogrel (out of patent at the time of writing) to alternatives not necessarily out of patent are needed.

This is the largest single study of primary care patients prescribed clopidogrel, incorporating *CYP2C29* genetic data and longer (>1 year) follow-up of clinical outcomes. However, this study is not without limitations. UK Biobank participants tended to be healthier than the general population at baseline assessment (13), however, linked primary care data used follow-up analysis should be unaffected by sample response patterns. Population attributable risk may be underestimated, as UK Biobank has a limited age range (mean age of 64 years at first prescription, max 79) that does not include many older people at high risk of strokes or MIs, so estimates of absolute risk may be greater in the general population. Further work is required to model this in other countries. The availability of only primary care prescribing (with no hospital prescribing details available) may mean that some clopidogrel prescriptions have been missed and therefore some patients or exposure periods excluded, but likely only for periods during and soon after hospital admissions. Although this is a relatively large study of 7,483 patients, stratified analyses (such as *2 heterozygous/homozygous groups, or by aspirin co-prescribing) were underpowered. The analysis was restricted to UK Biobank participants of genetically European ancestry (93% of the cohort); only 492 non-European participants had sufficient clopidogrel data, but this included all other ancestries including South or East Asian, African, and mixed, so analysis was not possible. Future analyses with more data will be needed to address these research questions. Similarly, analyses of the low frequency variants in larger samples may be needed to refine estimates, including for rs187941554, for which our nominally significant association (HR 5.44: 95% CIs 1.33 to 22.30, p=0.019) was based on only two incident strokes in 32 subjects.

To conclude, intermediate and low metabolizers (due to *CYP2C19* loss of function alleles) who were prescribed clopidogrel in the primary care setting had substantially increased risks of incident hospital-diagnosed strokes and MIs, compared to normal metabolizers. Evidence for improved patient outcomes with *CYP2C19* genotype-guided antiplatelet therapy is increasing, especially in the acute setting. Genotype guided (or routine) prescription of antiplatelet medications unaffected by CYP2C19 variants may improve outcomes in primary care patients for whom clopidogrel is currently indicated.

## Data Availability

Data are available on application to the UK Biobank (www.ukbiobank.ac.uk/register-apply). Additional data are available from the corresponding author on reasonable request.

## Acknowledgements

Access to UK Biobank resource was granted under Application Number 14631. We would like to thank UK Biobank participants and coordinators for this dataset. The authors would like to acknowledge the use of the University of Exeter High-Performance Computing (HPC) facility in carrying out this work.

## Contributors

LCP performed the analysis, interpreted results, created the figures, searched literature, and co-wrote the manuscript. DT and HF performed the literature search, performed analyses, and contributed to the manuscript. JA and JD performed analyses and contributed to the manuscript. CK, GAK, LF and JB contributed to data interpretation and contributed to the manuscript. JM provided expert clinical interpretation of the data and contributed to the manuscript. DM oversaw data analysis, interpretation, and literature searching, and led the writing of the manuscript. DM is the guarantor. The corresponding author attests that all listed authors meet authorship criteria and that no others meeting the criteria have been omitted.

## Ethical approval

The North West Multi-Centre Research Ethics Committee approved the collection and use of UK Biobank data (Research Ethics Committee reference 11/NW/0382).

## Transparency

The lead author (DM) affirms that this manuscript is an honest, accurate, and transparent account of the study being reported; that no important aspects of the study have been omitted; and that any discrepancies from the study as planned (and, if relevant, registered) have been explained.

