## Supplementary Material for "Genetic variation in activating clopidogrel: longer-term outcomes in a large community cohort"

### Clopidogrel, *CYP2C19* genetic variation, and outcomes in UK Biobank

Pilling et al.

### Supplementary Material

### Supplementary Methods

#### Prescription data

To identify prescriptions of clopidogrel and aspirin we used the UK Biobank "Coding system lookups and mappings" file (https://biobank.ctsu.ox.ac.uk/crystal/refer.cgi?id=592). For clopidogrel we searched for "clopidogrel|plavix|grepid" to include other relevant drug names. For aspirin we searched for "aspirin|nu-seals|caprin|disprin" also including relevant alternate names. We then used the associated read 2 codes and drug names in the `gp_scripts` UK Biobank prescription data for the participants. We did not use BNF codes as the resolution was not great enough to uniquely identify individual drugs.

### Supplementary Table 1: prescription codes in UK Biobank

| medication | term_description | read_2 | bnf_code |
|---|---|---|---|
| clopidogrel | clopidogrel | bu5.. | 02.09.00.00 |
| | clopidogrel 300mg tablets | bu54. | 02.09.00.00 |
| | clopidogrel 75mg tablets | bu51. | 02.09.00.00 |
| | grepid 75mg tablets | bu55. | 02.09.00.00 |
| | plavix 300mg tablets | bu53. | 02.09.00.00 |
| | plavix 75mg tablets | bu52. | 02.09.00.00 |
| aspirin | *aspirin 100mg m/r tablets | bu29. | 02.09.00.00 |
| | *aspirin 300mg m/r tablets | bu2b. | 02.09.00.00 |
| | *aspirin 324mg e/c tablets | di1h. | 04.07.01.00 |
| | *aspirin 500mg m/r tablets | di19. | |
| | *aspirin 600mg e/c tablets | di1g. | 04.07.01.00 |
| | *aspirin 600mg tablets | di1i. | 00.00.00.00 |
| | *aspirin 75mg tablets | bu25. | 02.09.00.00 |
| | *caprin 300mg e/c tablets | di1k. | 04.07.01.00 |
| | *caprin 324mg e/c tablets | di1a. | 04.07.01.00 |
| | *caprin 75mg e/c tablets | bu2F. | 02.09.00.00 |
| | *disprin cv 100mg m/r tablets | bu28. | 02.09.00.00 |
| | *disprin cv 300mg m/r tablets | bu2a. | 02.09.00.00 |
| | aspirin 100mg effervescent tablets | bu21. | 02.09.00.00 |
| | aspirin 150mg suppositories | di1o. | 04.07.01.00 |
| | aspirin 162.5mg m/r capsules | bu2I. | 02.09.00.00 |
| | aspirin 300mg dispersible tablets | j112. | 10.01.01.00 |
| | aspirin 300mg e/c tablets | di1f. | 04.07.01.00 |
| | aspirin 300mg effervescent tablets | bu27. | 02.09.00.00 |
| | aspirin 300mg soluble tablets | di1m. | 04.07.01.00 |
| | aspirin 300mg suppositories | di1n. | 04.07.01.00 |
| | aspirin 300mg tablets | j111. | 10.01.01.00 |
| | aspirin 75mg dispersible tablets | di13. | |
| | aspirin 75mg dispersible tablets | bu23. | 02.09.00.00 |
| | aspirin 75mg e/c tablets | bu2B. | 02.09.00.00 |
| | aspirin 75mg soluble tablets | bu2c. | 02.09.00.00 |
| | aspirin [antiplatelet] | bu2.. | 02.09.00.00 |
| | aspirin [central nervous system use] | di1.. | 04.07.01.00 |
| | aspirin [cns] 300mg dispersible tablets | di12. | 04.07.01.00 |
| | aspirin [cns] 300mg tablets | di11. | 04.07.01.00 |
| | aspirin [musculoskeletal use] | j11.. | 10.01.01.00 |
| | aspirin and the salicylates | j1... | 10.01.01.00 |
| | aspirin+metoclopramide 900mg/10mg/sachet powder | dl1b. | 04.07.04.01 |
| | aspirin+papaveretum 500mg/7.71mg dispersible tablets | diaG. | 04.07.01.00 |
| | aspirin/paracetamol/codeine tablets | dia1. | 04.07.01.00 |
| | dipyridamole+aspirin | bu4.. | 02.09.00.00 |
| | dipyridamole+aspirin 200mg/25mg m/r capsules | bu41. | 02.09.00.00 |
| | disprin 300mg dispersible tablets | di1r. | 04.07.01.00 |
| | isosorbide mononitrate+aspirin | blm.. | 02.06.01.00 |
| | isosorbide mononitrate+aspirin 60mg/150mg m/r tablets | blmy. | 02.06.01.00 |
| | isosorbide mononitrate+aspirin 60mg/75mg m/r tablets | blmz. | 02.06.01.00 |
| | nu-seals aspirin 300mg e/c tablets | di1c. | 04.07.01.00 |
| | nu-seals aspirin 600mg e/c tablets | di1d. | 04.07.01.00 |
| | nu-seals aspirin 75mg e/c tablets | bu2A. | 02.09.00.00 |
| | nu-seals cardio 75 e/c tablets | bu2G. | 02.09.00.00 |

**Supplementary Table 2: *CYP2C19* genotypes in UK Biobank**

| | | | | | | | UK Biobank | | | | | GnomAD | |
|---|---|---|---|---|---|---|---|---|---|---|---|---|---|
| CYP2C19 | Function | RSID | CHR | BP | A1 | A2 | HWE_p | INFO | MAF1 | MAF1 % | note | MAF2 | MAF2 % |
| *2 | LoF | rs4244285 | 10 | 96541616 | G | A | 0.9067 | 0.9998 | 0.1492 | 14.924 | Imputed | 0.1468 | 14.6800 |
| *3 | LoF | rs4986893 | 10 | 96540410 | G | A | 1.0000 | 1.0000 | 0.0001 | 0.006 | Directly genotyped | 0.0003 | 0.0264 |
| *4 | LoF | rs28399504 | 10 | 96522463 | A | G | 0.1615 | 0.7339 | 0.0022 | 0.216 | Imputed | 0.0025 | 0.2528 |
| *5 | LoF | rs56337013 | 10 | 96612495 | C | T | | | | | Not in UKB imputed | 0.0000 | 0.0008 |
| *6 | LoF | rs72552267 | 10 | 96535210 | G | A | | | | | Not in UKB imputed | 0.0003 | 0.0333 |
| *7 | LoF | rs72558186 | 10 | 96541756 | T | C | | | | | Not in UKB imputed | 0.0000 | 0.0000 |
| *8 | LoF | rs41291556 | 10 | 96535173 | T | C | 0.0415 | 1.0000 | 0.0030 | 0.304 | Directly genotyped | 0.0027 | 0.2710 |
| *17 | GoF | rs12248560 | 10 | 96521657 | C | T | 0.7279 | 0.9984 | 0.2154 | 21.539 | Imputed | 0.2314 | 23.1400 |

Function = LoF (Loss of Function, poor metaboliser), GoF (Gain of Function, rapid metaboliser)
BP = base position (hg19, build 37)
A1 = common allele
A2 = minor allele
HWE_p = Hardy-Weinberg deviation p-value
INFO = imputation quality score
MAF1 = minor allele frequency in UK Biobank Europeans
MAF2 = GnomAD European (non-Finnish) population minor allele frequency. GnomAD data can be viewed using URLs https://gnomad.broadinstitute.org/variant/rs4244285 and substituting the RSID for the appropriate variant.

### Supplementary Table 3: *CYP2C19* associations with incident outcomes stratified by *17 genotype

| Outcome / CYP2C19 genotype | N | N cases | Person-years | HR | 95% CIs | | *p* |
|---|---|---|---|---|---|---|---|
| ***Ischemic stroke*** | | | | | | | |
| Normal (*1/*1) | 2,948 | 37 | 7,208 | | | | |
| Intermediate/poor (any *2-*8) | 1,668 | 33 | 3,919 | 1.61 | 1.01 | 2.58 | 0.047 |
| Poor or rapid heterozygote (*2–*8/*17) | 476 | 8 | 1,191 | 1.25 | 0.58 | 2.69 | 0.570 |
| Rapid (any *17) | 2,385 | 32 | 6,039 | 0.99 | 0.62 | 1.59 | 0.970 |
| *Total* | *7,477* | *110* | *18,358* | | | | |
| ***Myocardial infarction*** | | | | | | | |
| Normal (*1/*1) | 2,905 | 707 | 5,403 | | | | |
| Intermediate/poor (any *2-*8) | 1,634 | 442 | 2,805 | 1.16 | 1.03 | 1.30 | 0.017 |
| Poor or rapid heterozygote (*2–*8/*17) | 466 | 112 | 904 | 0.98 | 0.81 | 1.20 | 0.880 |
| Rapid (any *17) | 2,354 | 562 | 4,691 | 0.95 | 0.85 | 1.06 | 0.330 |
| *Total* | *7,359* | *1823* | *13,803* | | | | |

*Analysis of European-ancestry participants with >1 clopidogrel prescription in the available GP prescribing data. Participants excluded if clopidogrel prescribing frequency was less than once every 2 months. Events <1 week after first clopidogrel prescription are excluded. Events occurring after the last known date of clopidogrel prescription are also excluded. HR=Hazard Ratios from Cox's proportional hazards regression models adjusted for age at first clopidogrel prescription, sex, and genetic principal components of ancestry 1-10. CI = Confidence Intervals.*
